# Polybrominated diphenyl ether serum concentrations and depressive symptomatology in pregnant African American women

**DOI:** 10.1101/2020.10.13.20212316

**Authors:** A. Mutic, D. Barr, V. Hertzberg, A. Dunlop, P. Brennan, L. McCauley

## Abstract

**Background:** Polybrominated diphenyl ethers (PBDEs) are lipophilic, persistent endocrine disrupting chemicals often used as flame retardants in products that were widely produced in the United States until 2004. The potential for environmental toxicants such as PBDEs to disrupt normal neuroendocrine pathways resulting in depression and other neurological symptoms has been largely understudied. This study examined whether PBDE exposure in pregnant women was associated with antenatal depressive symptomatology.

**Methods:** This study is part of a larger longitudinal pregnancy and birth cohort study. Data were collected from 193 African American pregnant women at 8-14 weeks gestation. Serum PBDEs were analyzed using gas chromatography-tandem mass spectrometry. The Edinburgh Depression Scale (EDS) was used to identify depressive symptoms experienced in the last seven days prior to biosampling. The dichotomous depression variable was used to explore varying high-risk EDS cutoffs and illustrated with receiver operating characteristic curves. Logistic regression models were constructed to investigate associations with antenatal depression and a weighted quantile sum (WQS) index was calculated to account for the mixture of PBDE congeners.

**Results:** Of the total sample, 52 women (26.9%) were categorized as having a high risk of depression. PBDE congeners −47, −99, and −100 were detected in 50% or more of the samples tested. BDE-47 was positively associated with depressive symptoms (β =2.36, p=0.05). The risk of being mild to moderately depressed increased by a factor of 4.52 for BDE-47 (CI 1.50, 13.60) and 1.58 for BDE-99 (CI 1.08, 2.29). The WQS index, a weighted estimate of the body burden of the congener mixture was positively associated with a higher risk of mild to moderate depression using an EDS cutoff ≥10 (OR=2.93; CI 1.18, 7.82).

**Conclusion:** BDE-47 and −99 exposures are significantly associated with depressive symptomatology in a pregnant cohort. These exposures will likely continue for years due to slow chemical degradation. Interventions should focus on PBDE mitigation to reduce toxic neuroendocrine effects on vulnerable pregnant women.

## BACKGROUND

Polybrominated diphenyl ethers (PBDEs) are a family of chemicals with a common structure of a diphenyl ether molecule attached to one to ten bromine atoms. PBDEs were once added to consumer products and materials to attenuate the risk of fire and to increase the escape time in the event of a fire. A voluntary phase-out of pentaBDEs and octaBDEs began in the United States in 2004 and later, decaBDEs, however, they remain prevalent in the environment over 10 years later. PBDE chemicals are not covalently bound to materials and leach from consumer products into the environment. Additionally, they are not easily biodegraded and thus remain persistent and ubiquitous in the environment. Levels of PBDEs in humans and the environment are higher in North America than in other regions of the world, likely because of the widespread commercial use of PBDE mixtures in the United States: decaBDE and pentaBDE (widely used in textiles, plastics, electronics, and polyurethane foam)(EPAb, 2006). The chemicals are highly detectable in air, dust, sediment, soil, and ground water (ATSDR, 2017) and, with their hydrophobic properties, PBDEs can bioaccumulate in lipid-rich human tissue (NTP, 2014). Exposure to PBDEs has been associated with thyroid disease (Vuong, 2018; Byrne, 2018; Chen, 2018), reproductive changes (Harley, 2010), neurodevelopmental deficits (Kuriyama, 2005; Shy et al., 2011; Hoffman et al., 2012; Bradman et al., 2012; Eskenazi et al., 2011, 2013), and gestational diabetes (Liu, 2018). The potential for environmental toxicants such as PBDEs to disrupt normal neuroendocrine pathways resulting in depression and other neurological symptoms has been largely unstudied. Literature regarding PBDE exposure and subsequent short- and long-term health effects has focused on neurological effects among infants and children. To date, only <u>two studies</u> have investigated the effects of PBDE exposure outside of pregnancy *and* subsequent neuropsychological effects (Gump, 2014; Fitzgerald, 2012). In this study, we focus on PBDE exposure in pregnant adults and their relationship with depressive symptoms.

Clear connections between PBDE levels and thyroid disruption have been made (Vuong, 2018; Guo, 2018; Byrne, 2018; Chen 2018; Ding, 2017; Zheng, 2017) PBDEs have similar shape and structure to thyroid hormones and are thought to mimic thyroid activities as a result. Further, the essentiality of thyroid homeostasis for cognition and mood stability has been long recognized (Hendrick, 1998; Joffee, 1994; Esposito, 1997; Bathla, 2016). Of the two available studies investigating PBDE exposure and risk of depression within the same individual, no associations were found. However, measures of hostility, aggression, and temperament among children were significantly correlated with PBDE concentrations (Gump, 2014). In another study of older adults with high occupational polychlorinated biphenyl (PCB) exposure, significant associations were found between the mixture of PCBs and PBDEs and learning and memory (Fitzgerald, 2012). PCBs and PBDEs have been postulated to share mechanistic actions and toxic effects (Sethi 2017, Herbstman 2010; Dingemans 2016; Harley 2010) and others endocrine disrupting chemicals and have been correlated with depression-like symptoms as well (Fitzgerald, 2007; Fitzgerald, 2012; Xu, 2015; Braun, 2011; Engel, 2010).

Pregnant women may be an overlooked vulnerable population to the effects of toxicants due to normal physiological changes in pregnancy such as higher lipid profiles and thyroid strain (Lignell, 2016; Tan, 2013; Budenhofer, 2013). African Americans (AAs) specifically may also be at elevated risk of toxicant exposure and depression. According to the most recent 2003-2008 National Health and Nutrition Examination Survey (NHANES) population data, non-Hispanic blacks have elevated PBDE body burden when compared to non-Hispanic whites (Sjodin, 2008; Sjodin, 2014). Cross-sectional studies have found similar findings predicting non-Hispanic blacks are nine times more likely to have high concentrations of persistent organic pollutants (which include PBDEs) than non-Hispanic whites after adjusting for income (Pumarega, 2016).

AA women may also be at increased risk for perinatal depression-a mood disorder present in the antenatal or postpartum period for up to one year. Melville et al. (2010) estimated nearly 19% of AA women met diagnostic criteria for either minor or major depression during pregnancy, considerably higher than the national average of 8.5% (Tanden, 2012). African American women of low income have the highest reported prevalence of perinatal depression (28%) in the U.S. (CDC). Major depressive disorder (MDD) is associated with adverse outcomes in many aspects of role performance (Kessler 2012). Specifically, MDD is associated with poor outcomes in marital functioning (cite), parental functioning (cite), absenteeism and low work performance (cite), and personal income or household earnings (cite). Unfortunately, some populations of African Americans are less likely to be diagnosed and treated for depressive symptoms compared to white patients (Gallo et al., 2005), and across subgroups, African Americans are more likely to have persistent disorders after becoming ill (Breslau 2005). Additionally, mental health concerns are often overlooked, misdiagnosed, mistreated, or not treated and, historically, both patients and providers have poor follow-up of mental health concerns.

It is also widely accepted that not all populations conform to the standard EDS scoring criteria. The EDS has been validated in multiple race and ethnic groups (Kozinszky, 2015, Cox, 1987) with varied cut-off scores recommended to accurately predict a diagnosis of mild to moderate depression resulting from socioeconomic status (Tanden, 2012), race (Tanden, 2012), and gender (Matthey, 2001) differences. Additionally, researchers have argued that cultural differences and timing of screenings influence clinical cut-off scores (Tandon 2012, Chaudron et al., 2010; Gjerdingen et al., 2009; Sheeder et al., 2009; Logsdon & Myers, 2010).

Few quantitative studies have examined health effects of PBDE exposures in pregnant women and, to our knowledge, <u>none</u> have studied AA women. For these reasons, we designed our study to investigate whether PBDE exposure in pregnant AA women is associated with depressive symptomatology during pregnancy. Due to uncertainty in applicability of standard EDS cutoff scores to our vulnerable population of pregnant AA women, we took additional measures to ensure appropriate cutoff scores were utilized in our analysis. We hypothesized a positive relationship would be found between PBDE serum concentration and depression among pregnant AA women.

## METHODS

This study used data drawn from a larger longitudinal pregnancy and birth cohort aimed to investigate the maternal prenatal microbiome as a predictor of birth outcomes (Corwin, 2017). Women were enrolled in the study between 2014 and 2017 and were included if they self-identified as AA, had a singleton pregnancy, were 18-35 years of age, resided in the Atlanta Georgia area, and could comprehend and speak English. Women were recruited during their first trimester from prenatal clinics serving two local hospitals. Members of this SES-diverse cohort were individually informed of the study protocol and written consent was obtained. Participants were followed through delivery. For this analysis, a subset of 193 women were included if they had serum PBDE concentrations and complete EDS scores. All study procedures were approved and reviewed by the Institutional Review Board at Emory University.

### Measures

Pregnancy dating was based on the participant’s known last menstrual period or first trimester ultrasound if a discrepancy of ± 5-7 days exists per standard dating protocol (Corwin, 2017). Measures (questionnaire, clinical, and biological data) were collected in person at the initial visit or by electronic medical record.

#### Demographic data

Socio-demographic variables included age, marital status, years of education, and insurance status. The poverty/income ratio was determined from family size and household income data.

#### Clinical and Edinburgh Depression Scale Data

Health data extracted from EMR included height, weight, parity, past and current tobacco use, and alcohol or drug use in the first trimester of pregnancy. Depression data was collected by trained research coordinators at 8-14 weeks gestation. Depression was defined using the Edinburgh Depression Scale (EDS); a 10-item scale of depressive symptoms experienced in the last seven days (Cox, 1987). The EDS is a reliable tool to predict antenatal combined major or minor depression (ACOG) with psychometric properties of 64-87% sensitivity, 73-96% specificity, and 73% positive predictive value (Kozinszky, 2015). Individual question scores were summed and total scores ranged from 0-30 with higher scores associated with a higher risk of depression. Four cut-off scores were explored (≥7, ≥8, ≥9, and the universal standard ≥10) to determine the optimal cutoff value to detect depressive symptomatology in our AA study sample (Cox, 1987). Receiver operating characteristic curves were constructed to visualize and compare the specificity of the cutoff scores. An optimal EDS cutoff of ≥10, area under the curve 0.6867, was determined to differentiate the groups of high versus low or no risk of depression (Figure 1).

**Figure 1.**
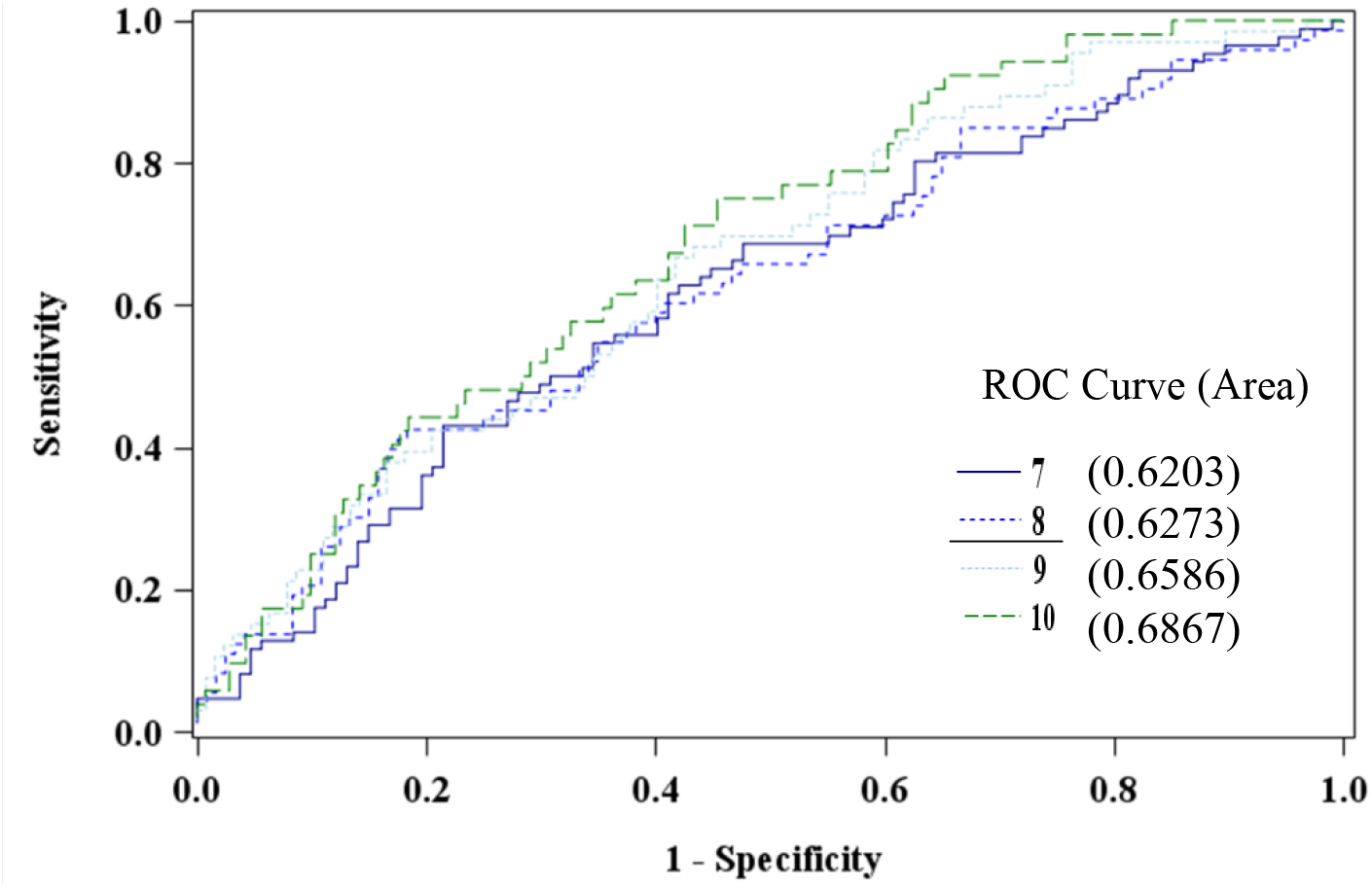
Comparison of Edinburgh Depression Scale Cut Points using PBDE 47.

#### Serum PBDE Concentrations

Concentrations of PBDEs congeners 47, 85, 99, 100, 153, and 154 were measured in maternal serum using a modification of a previous method (Darrow, 2017; Jacobson, 2016). Samples were fortified with isotopically labeled analogues of the target chemicals, homogenized and deprotonated. Supernatants were extracted twice with hexane and dichloromethane and passed through an activated silica gel column to remove residual biogenic material. Sample extracts were concentrated and analyzed using gas chromatography-tandem mass spectrometry with isotope dilution calibration. The limits of detection (LODs) (see supplemental Table 1) were in the low pg/mL range. Values below the LOD were replaced with a value equal to the LOD, divided by the square root of two (Hornung and Reed, 1990). Maternal serum total cholesterol and free triglycerides were not collected, therefore lipid adjustment was not done.

**Table 1.** Demographic characteristics of AA cohort 2014-2015 associated with high depressive symptoms using an EDS cutoff ≥10.

|  | Total sample<br>(n=193) | High depressive<br>symptoms<br>(n=52) | Low or no<br>depressive<br>symptoms | P value |
| --- | --- | --- | --- | --- |

|  |  |  | (n=141) |  |
| --- | --- | --- | --- | --- |
| Characteristics | n (%) | n (%) | n (%) |  |
| Age (mean, SD) | 24.0 ± 4.4 | 23.8 ± 3.9 | 24.4 ± 4.6 | .46 |
| Married | 26 (13.5) | 3 (5.8) | 23 (16.3) | .06 |
| Education |  |  |  | .30 |
| Some high school | 35 (18.1) | 12 (23.1) | 23 (16.3) |  |
| Graduated high school or GED | 65 (33.7) | 18 (34.6) | 47 (33.3) |  |
| Some college or technical school | 63 (32.6) | 16 (30.8) | 47 (33.3) |  |
| Graduated college | 20 (10.4) | 6 (11.5) | 14 (9.9) |  |
| Some graduate work or degree | 10 (5.2) | 0 (0.0) | 10 (7.1) |  |
| Insurance |  |  |  | .08 |
| Low Income Medicaid<100% | 53 (27.5) | 10 (19.2) | 43 (30.5) |  |
| Right from the Start Medicaid≤ 200% | 97 (50.3) | 33 (63.5) | 64 (45.4) |  |
| Private | 43 (22.2) | 9 (17.3) | 34 (24.1) |  |

### Covariates and Potential Confounders

In order to test the relationship between PBDE congeners and depression, covariates and/or potential confounders were considered *a priori* and reassessed after bivariate analyses. The confounders identified *a priori* were age, gravidity, marital status, education, BMI, and income based on associations noted in previous literature (Faravelli, 2013; Feder, 2018; Bracke, 2014; Wiltink, 2013). Income had >20% missing values (n = 39) so insurance type was used as a proxy. Parity, indicating the number of pregnancies reaching viable gestational age, was tested as a covariate. During bivariate analysis, education and parity were dropped because no significant associations with the outcome of depression were observed.

### Statistical Analysis

PBDE congeners were log transformed (base 10) to reduce the influence of outliers and to conform to normality. For congeners with a 50% or greater presence (−47, −99, and −100), geometric means were used to describe PBDE data and reduce the right skewed data distribution resulting from imputed data. Congeners −85 and −154 were not detected in any samples (above instrumental LOD) and were therefore not reported and excluded from analysis. Congener −153 was detected in only 14% of the samples and was subsequently not used in further analyses.

We examined univariate associations within demographic characteristics, PBDE concentrations, and EDS scores. Geometric means were used to describe PBDE concentrations and to compare to 2013-2014 NHANES data restricted to non-Hispanic black women aged 18-35 years. Wilcoxon nonparametric tests were used to compare PBDE congeners between low and high depression groups. Parametric tests of association were completed for covariates and depression variables. Spearman rank-order and Kruskal-Wallis tests were conducted to compare means among skewed data. Bivariate analysis between PBDE 47 and depression groups yielded unequal variances so Satterthwaite test statistic was reported. Missing data were explored within each covariate and accounted for <10% of the sample and were not further manipulated.

Each PBDE congener was regressed with confounding variables in linear and logistic regression models to investigate associations with depression. Exposure to a PBDE mixture (congeners −47, −99, and −100) was evaluated using a weighted quantile sum (WQS) approach in conjunction with multiple linear and nonlinear logistic regression (Carrico, 2014). The WQS method estimates a weighted linear index corresponding to chosen quantiles of PBDEs. Bootstrap sampling is used to empirically determine the weights, constrained between 0 and 1 and summed to 1. Since environmental exposures co-occur, interact, and are highly correlated, the WQS method reduces dimensionality and addresses multicollinearity (Czarnota 2015). The WQS index was placed in the best-fit model where exp(β1) is the odds ratio associated with a unit (quartile) increase in the weighted quartile sum of PBDE exposures. The WQS index was regressed using an EDS cutoff ≥10. The significance of the test represents a test for a mixture effect.

Statistical significance was set at 0.05 and all analyses were performed using SAS v9.4 and R v3.4.

## RESULTS

Basic descriptive statistics for the total sample of 193 women are presented in Table 1. Of the total sample, 52 women (26.9%) were categorized as having a high risk of depression (EDS cutoff ≥10) and 141 (73.1%) had a low or no risk of depression. In this sample, the alpha coefficient was 0.85. The majority of participants were unmarried (86.5%), had at least some college education (48.2%), and qualified for Medicaid health insurance (77.8%) (Table 1). Both high and low or no risk of depression groups had an approximately normal distribution of educational attainment. Approximately half were overweight or obese (54.4%) and nulliparous (48.2%). A small proportion reported consuming alcohol (4.2%) or marijuana (17.1%) in the last month, or ever smoked tobacco (10.9%) (Table 2).

**Table 2.** Health-related characteristics of AA cohort 2014-2015 associated with high depressive symptoms using an EDS cutoff of ≥10.

|  | Total sample<br>(n=193) | High depressive<br>symptoms<br>(n=52) | Low or no<br>depressive<br>symptoms<br>(n=141) | P value |
| --- | --- | --- | --- | --- |
| Characteristics | n (%) | n (%) | n (%) |  |
| Body mass index |  |  |  | .03* |
| Underweight (<18.5) | 8 (4.2) | 4 (7.7) | 4 (2.8) |  |
| Normal (18.5 – < 25) | 80 (41.5) | 24 (46.2) | 56 (39.7) |  |
| Overweight (25 – < 30) | 39 (20.2) | 14 (26.9) | 25 (17.7) |  |
| Obese ($\geq 30$) | 66 (34.2) | 10 (19.2) | 56 (39.7) | |
| Number of children |  |  |  | .63 |
| 0 | 93 (48.2) | 28 (53.9) | 65 (46.1) |  |
| 1 | 58 (30.1) | 14 (26.9) | 44 (31.2) |  |
| 2+ | 42 (21.8) | 10 (19.2) | 32 (22.7) |  |
| Drinks alcohol in last month | 8 (4.2) | 4 (50.0) | 4 (50.0) | .14 |
| Ever smoked tobacco | 21 (10.9) | 14 (26.9) | 7 (5.0) | <.0001* |
| Smoked marijuana in last month | 33 (17.1) | 14 (26.9) | 19 (13.5) | .03* |

\*Statistical significance at $p < 0.05$

The women’s mean BMI scores were significantly different between the depression groups. Substance use in the last month and ever used tobacco were also associated with having an EDS score of ≥10.

When comparing the 2014-2014 NHANES data reflecting the pooled samples of non-Hispanic Black women aged 20-39, our study population had similar levels of BDE −47 and −99 but lower levels of BDE −100 (Table 3).

**Table 3.** Wet weight PBDE congeners present in the study population and compared to 2013-2014 NHANES data reported in pg/mL serum (N=193).

| Metabolite | mean ± SD | GM | NHANES mean | % detected |
| --- | --- | --- | --- | --- |
| **PBDE 47** | 124.9 ± 137.6 | 86.8 | 87.3 | 100% |
| **PBDE 99** | 35.0 ± 45.3 | 20.8 | 18.3 | 81% |
| **PBDE 100** | 22.3 ± 21.1 | 13.4 | 18.2 | 79% |

\* Reported NHANES data reflects arithmetic mean from pooled samples of non-Hispanic Black women aged 20-39 years.

Consistent with previous literature, PBDE congeners were significantly intercorrelated (r= 0.6-0.7, p<0.001), limiting statistical methods. Although not strong, serum PBDE 47 concentrations and marijuana use were positively associated (r=0.12, p=0.03). Neither tobacco nor alcohol use were associated with any congener.

Levels of BDE −47 and −99 were significantly different between low and high depression groups (p=0.04 and p=0.02, respectively) (Table 4). As concentrations of PBDE −47 in the serum increase, the probability of having depressive symptoms also increases (β =2.36, p=0.05) (Figure 2). However, this association accounted for only 6% of the variability.

**Table 4.** PBDE concentrations (pg/mL) in serum and risk of antenatal depression among African American cohort at 8-14 weeks gestation (N=193).

|  | **High depressive symptoms**<br>**(n=52)** | **Low or no depressive symptoms**<br>**(n=141)** |  |
| --- | --- | --- | --- |
| **Characteristic** | **mean ± SD** | **mean ± SD** | **P value** |
| PBDE 47 | 155.3 ± 198.3 | 113.7 ± 105.8 | .04* |
| PBDE 99 | 112.1 ± 342.9 | 91.4 ± 342.9 | .02* |
| PBDE 100 | 104.7 ± 343.0 | 94.1 ± 343.0 | .24 |

\*Statistical significance at p<0.05

**Figure 2.**
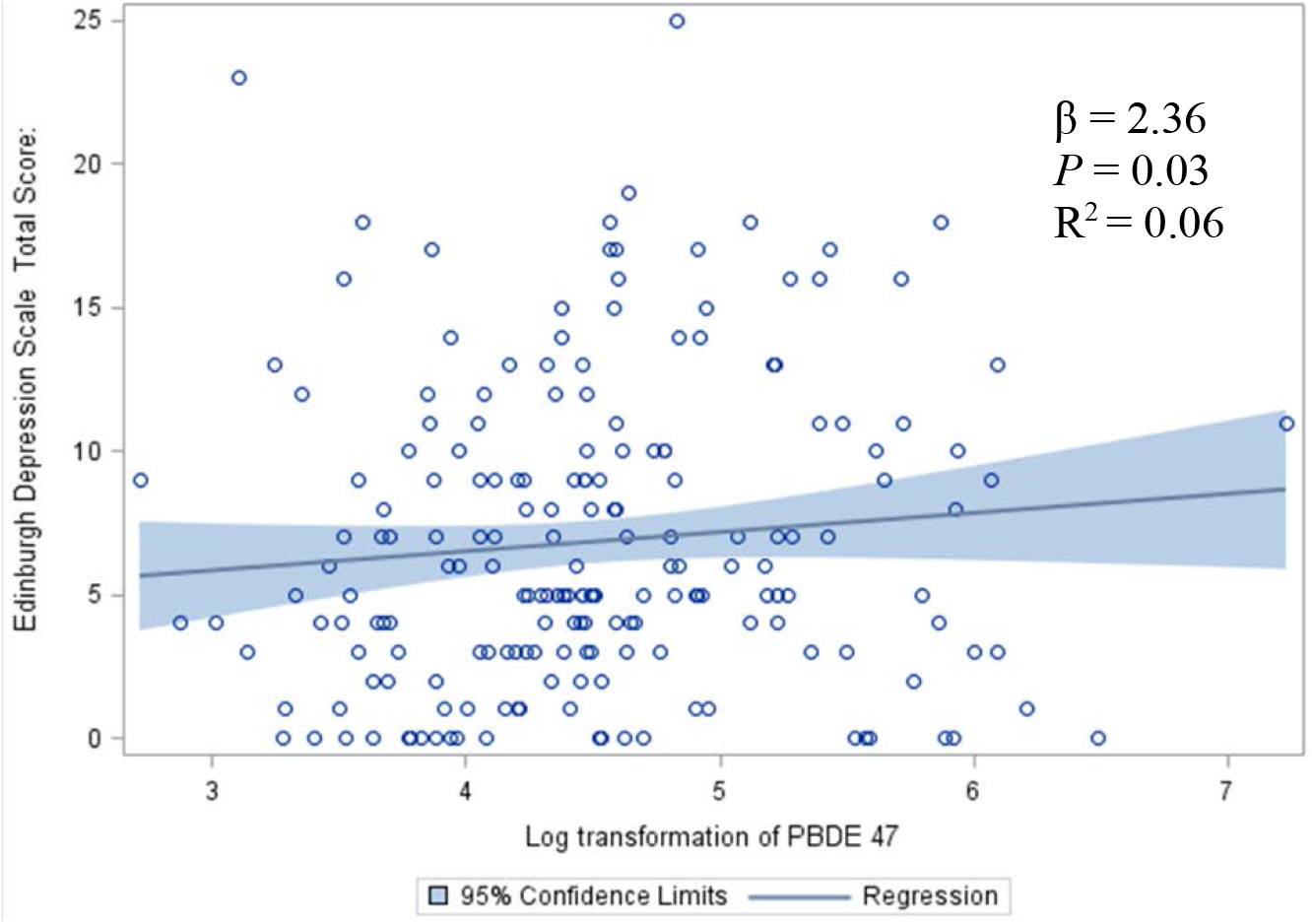
Relationship between PBDE47 and Depressive Symptoms.

All covariates and confounding variables identified *a priori* were entered into the multivariate model. The best-fit model controlled for age, marital status, and BMI to test for associations between PBDEs and depression.

In the adjusted regression analysis, models were used to examine the associations between PBDEs (Table 5), and depressive symptoms. Statistical significance was detected with BDEs −47 and 99, but not BDE −100. For every one unit increase in concentration, the risk of being mild to moderately depressed increased by a factor of

**Table 5.** Multiple logistic regressions of high antenatal depressive symptoms among African American women using an EDS cutoff ≥10.

|  | OR [95% CI] | AOR [95% CI] |
| --- | --- | --- |
| **PBDE 47** | 2.62 (1.00, 6.87) | 4.52 (1.50, 13.60)* |
| **PBDE 99** | 1.39 (0.99, 1.93) | 1.58 (1.08, 2.29)* |
| **PBDE 100** | 1.17 (0.88, 1.58) | 1.22 (0.90, 1.67) |

\*Statistical significance at $p < 0.05$
\*Adjusted for age, BMI, and marital status

4.52 for BDE-47, 1.58 for BDE-99, and 1.22 for BDE-100 adjusting for age, marital status, and BMI. No issues of multicollinearity existed since all various inflation factors values were low (<2). The Hosmer-Lemeshow goodness of fit (0.461-0.594) suggests good fit for each model.

In order to show which PBDE congener had the most robust association with depressive symptoms, a PBDE WQS index was regressed with the dichotomous depression variable and presented in Table 6. The distribution of weights comprising the index was led by BDE-47 (*w*= 0.71) followed by BDE-100 (*w*= 0.29), then BDE-99 (*w*= 0. 00000000817). The fitted coefficients exp(β) provided an estimated odds ratio for risk of depression resulting from the PBDE mixture. Specifically, increases in the weighted PBDE index were significantly associated with higher depressive symptoms after adjusting for age, marital status, and BMI (OR=2.93; CI 1.18, 7.82).

**Table 6.**
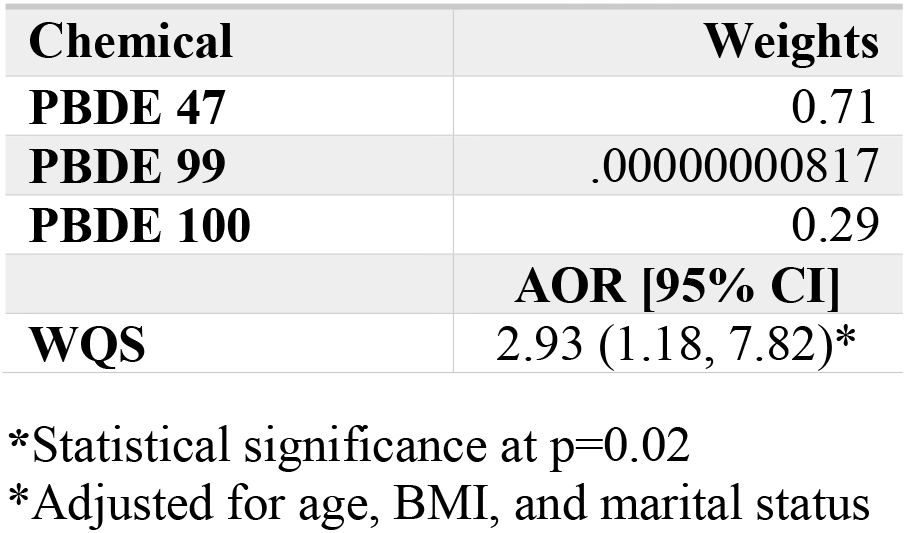
Associations between Weighted Quantile Sum Regression Index and high antenatal depression symptoms using an EDS cutoff ≥10 in the study population.

## DISCUSSION

The purpose of this study was to examine the relationship between concentrations of PBDEs in serum and depressive symptoms in pregnant AA women, a vulnerable and traditionally understudied population in the context of both PBDE exposure and prenatal mental health outcomes. The bulk of PBDE and health outcome research has centered on maternal exposures and fetal development but, as concentrations continue to be detect remote from the production phase-out, health concerns beyond early childhood should be explored. Cowell et al. (2018) found no significant differences in PBDE cord blood concentrations collected before and after the phase out supporting the persistent properties and ongoing release if the chemicals from products. Exposures and their body burdens are not well-characterized in pregnant women versus nonpregnant. In our sample of 193 pregnant women, 100% had detectable concentrations of BDE −47. Body burden estimates are similar to serum concentrations from the most recent 2013-2014 NHANES national sample of non-Hispanic black women of reproductive ages. Findings indicate positive associations between PBDE congeners, the PBDE mixture, and high depressive symptoms among our study population. The weighted PBDE index identified BDE −47 to be the “bad actor” within the mixture. We anticipated this since it is highly prevalent across multiple matrixes (cite). Due to the small amount of variability in the final regression models, further studies should focus on potential covariates that could better explain the relationship between serum PBDE concentration and prevalence of depressive symptoms. We understand humans are exposed to many environmental chemicals at once and the preliminary findings in this study using the WQS method broadens our perspectives on analyzing highly correlated variables and provides better estimates of the PBDE mixture effect. For more comprehensive mixture estimates, additional endocrine disrupting chemicals should be considered. Theoretically, endocrine disruptors have similar health effects and may be working through similar pathways (cite) to affect mood. Assessing for exposure to other endocrine disruptors and neuropsychological outcomes in this population could provide insight into shared metabolic pathways.

Consistent with previous studies of tobacco use and depression in pregnancy (Newport, 2012), tobacco use and substance abuse correlated with EDS scores. However, our data collection protocol did not allow us to account for the onset of depressive symptoms to further analyze the correlations. It is plausible that depressed or stressed women are likely to smoke tobacco or use marijuana as a coping mechanism (Lobel, 2008; Weaver, 2008). It is also plausible there is no causal relationship between substance use or smoking and depression, but rather the behavior is an expression of one’s attitude or emotion (Rodell, 2009). Lazarus and Folkman (1987) described stress as being physiologic and psychologic, both having the opportunity to lead to problematic health outcomes. In the context of this study, we believe stressors lead to physiologic and psychologic depressive responses without regard to co-occurring behaviors such as smoking or marijuana use.

Based on this study, there is a high risk of perinatal depression among African American (27%) compared to the national average of all perinatal women regardless of race (8.5%). In our study of only AA women, those with lower income, using insurance as a proxy, had disproportionate rates of depression (29%) compared to those with higher income or private insurance (21%). This is consistent with other studies who have been reported elevated perinatal major depression rates of 19-28% (Melville 2010; Hobfall, 1995; Grobman, 2016; Tandon, 2012) also in low income AA women.

Our outcome measure, EDS score, is a measure of depression *risk*, not a diagnostic measure. It is primarily utilized by health professionals and researchers to objectively identify mothers suffering from stress that may be interfering with normal activities or enjoyment of life. Clinically, the EDS cutoff used for referral or to warrant repeat testing remains variable (Bunevicius, 2009; Tsai, 2013; Bergink, 2011). In light of this uncertainty, we took a careful analytic approach to identify an appropriate value specific for our sample. Surprisingly, our analysis demonstrates that using a standard cutoff of ≥10 is optimal for decision making in regards to depression risk for urban AA pregnant women. Using a cutoff of ≥10 is likely to accurately predict depression in most women but some suggest repeat testing as frequent as two weeks later regardless of the cutoff score (Matthey, 2012), especially among populations where cultural variations may exist in the expression of depressive symptoms (Smith-Nielson, 2018). Tandon et al. (2012) collected depression measures and performed subsequent sensitivities on the EDS. Similar to this study, they recommended an optimal cutoff score for major and minor depression of ≥10 in this population. In general, AAs are less likely to be diagnosed with depression or treated for depressive symptoms compared to white patients (Gallo et al., 2005), and across subgroups, African Americans are more likely to have persistent mental disorders after becoming ill (Breslau 2005). At a minimum, providers should be alerted when EDS scores fall between 7-10 since patients could be developing depression and a trustworthy rapport can positively impact patient compliance and follow-up (Zink 2017; Gilbert 2009).

### Limitations

There are notable limitations to this study. Since our study relied on maternal-self report to document tobacco use, substance abuse, and depressive symptoms, it is possible that recall bias interfered with our results. The study sample is based on convenience sampling from two metropolitan prenatal clinics. While it may represent most AA pregnant women living in urban environments, attempts were not made to include hard-to-reach groups. Especially during pregnancy, study participants may have been reluctant to disclose the presence or absence of depressive symptoms or current use of tobacco or illegal drugs. Women report difficulty disclosing perinatal mental illness because of stigmas of inadequacy as a mother or stigmas of treatment in pregnancy (Moore, 2016). Others refuse to seek help because of poor mental health knowledge or literacy (Jorm, 2006). Self-report of substance abuse is underrepresented resulting from maternal guilt or remorse (Stengel, 2014; Stone, 2015). A cross-sectional study design comprised of one time point, limits the ability to cultivate causal relationships. We attempted to compensate for this lack of temporality in part by excluding anyone with a personal or family history of depression. Further longitudinal data is needed to substantiate the findings and more confidently predict conclusions. Another limitation of our analytic approach was the lack of serum lipid data which hindered our ability to lipid adjust serum PBDE concentrations. Variation of PBDE concentrations in humans is common due to multiple exposure routes and elimination differences among each congener. A final key limitation was the inability to determine the major sources of environmental exposures to PBDEs in this population which impacts future development of preventive interventions for pregnant AA populations.

## CONCLUSION

Health concerns related to PBDE exposure are becoming apparent and will likely continue for years due to slow chemical degradation leading to persistence and accumulation in our environment. Products with PBDEs are common and continue to be used in homes, schools, and businesses across the country. Even those removed from homes continue to decompose in landfills and recycling centers releasing harmful toxicants into the air, water, and soil. Our analysis revealed BDE-47, −99, and −100 are present in a cohort of pregnant AA women and the weighted mixture is associated with high depressive symptoms. The novel associations were found in a relatively small sample size and among a non-occupationally exposed group. The need for replication and consistent results is suggested. Because of limited research and slow PBDE degradation, it is unclear whether PBDE accumulation has other long-term implications on pregnant women. Additional studies are needed to address population health impacts resulting from PBDE bioaccumulation since the chemicals are likely to be present in the environment for many years ahead. The cross-sectional methodology of this study can be particularly useful in informing researchers and public health professionals about depressive symptomatology among AA pregnant women and changes in PBDE concentrations over time. Assessing environmental burdens and human toxicant effects, especially mixtures, can inform planning and allocation of health resources for mitigation and prevention. Endocrine disrupting chemicals such as PBDEs are completely modifiable and prevention efforts should be given high priority.

## Data Availability

Data is not publicly available.

---

**SUPPLEMENTAL TABLE 1.**
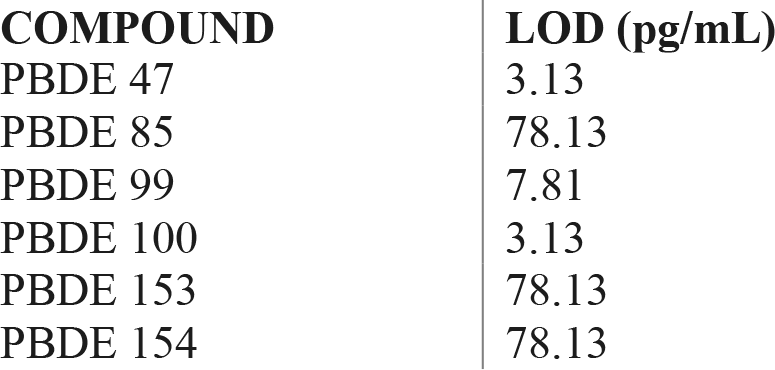
LEVEL OF DETECTION FOR EACH PBDE CONGENER.

| COMPOUND | LOD (pg/mL) |
| --- | --- |
| PBDE 47 | 3.13 |
| PBDE 85 | 78.13 |
| PBDE 99 | 7.81 |
| PBDE 100 | 3.13 |
| PBDE 153 | 78.13 |
| PBDE 154 | 78.13 |

**Supplemental Figure 1.**
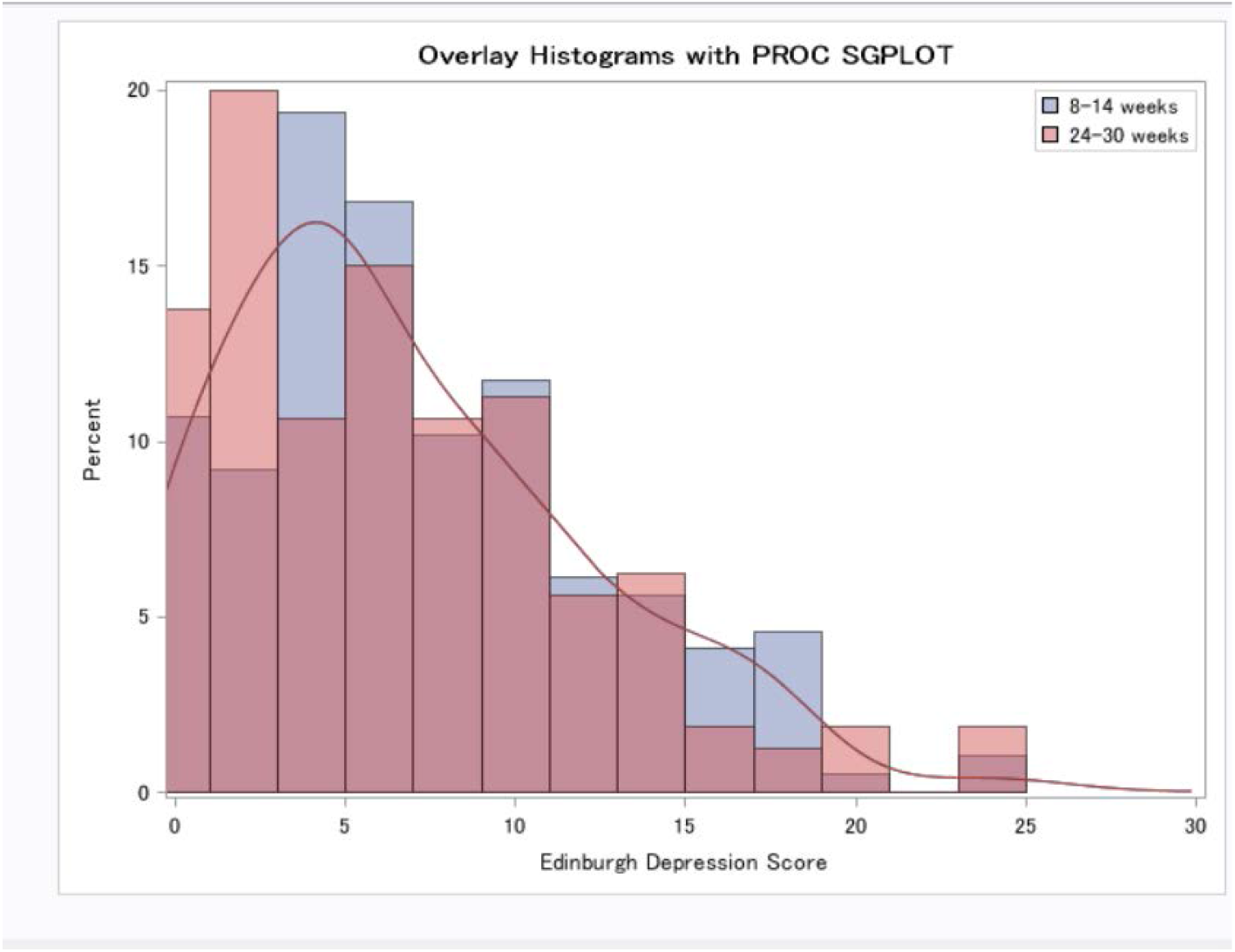
Comparison of EDS scores between 8-14 weeks and 24-30 weeks gestation.

**Supplemental Table 2.** Comparison of high depressive symptoms using an EDS cutoff ≥10 at 8-14 weeks and 24-30 weeks gestation (N=193).

| Time point | N | EDS Score Mean (SD) | EDS Score $\geq 10$ N (%) | Difference in Mean P Value |
| --- | --- | --- | --- | --- |
| 8-14 weeks | 193 | 6.82 (5.24) | 52 (27) | 0.28 |
| 24-30 weeks | 160 | 6.20 (5.52) | 39 (20) |  |

There was no significant difference noted in depression between the two time points 8-14 weeks and 24-30 weeks gestation (Supplemental Table 1).

## Notes

### Competing Interest Statement

The authors have declared no competing interest.

### Funding Statement

 
This study was funded by a grant from the National Institutes of Health, National Institute of Nursing Research (R01NR014800) as well as the National Institute of Environmental Health Sciences (P50ES026071) and US EPA (83615301).

### Author Declarations

Emory University Institutional Review Board

